# An optimal control policy for COVID-19 pandemic until a vaccine deployment

**DOI:** 10.1101/2020.09.26.20202325

**Authors:** Hamid R. Sayarshad

## Abstract

When an outbreak starts spreading, policy makers have to make decisions that affects health of their citizens and the economic. Some might induce harsh measures, such as lockdown. Following a long harsh lockdown, economical declines force policy makers to rethink reopening. But what is the most effective reopening strategy? In order to provide an effective strategy, here we propose a control strategy model. Our model assesses the trade-off between social performance and limited medical resources by determining individuals’ propensities. The proposed strategy also helps decision makers to find optimal lockdown and exit strategy for each region. Moreover, the financial loss is minimized. In addition, a study of a COVID-19 dataset for Los Angeles County is performed to validate our model and its results.

## 1 Introduction

According to World Health Organization (WHO) the first death due to SARS-COV-2 infection was reported on January 11, 2020 in China Wuhan [1]. The origin of the virus still remains uncertain. By the end of August 2020, close to 25 million people had been infected worldwide by COVID-19 virus and over 843,000 people have died (John Hopkins Center [2]). The virus spread between nations quickly and it became a global concern.

Currently many countries are practicing community-based measurements to mitigate the spread of the virus. The policy makers and governments have tried to control the epidemic based on the limited medical resources that makes a delay in the infection peak, providing more time to produce a vaccine [3]. As a result, the closure of many businesses has been implemented which leads to some economical loss [4]. It raises the question: “what are the alternative interventions with lowest negative economic impact?”

When a *Susceptible* person makes a contact with a contagious individual, he/she may become *Exposed*. It will take some time till the symptoms appear. A person might become *Infectious* before being symptomatic. Finally that person will be *Removed* either through death or recovery with temporary immunity. SEIR models or SIR models, as their names indicate are mathematical models that are used to describe individuals transitioning between these stages. For classical references on virus dynamics refer to [5, 6, 7, 8].

To address several questions on the control strategy of the COVID-19 pandemic, we propose an optimization model based on the SIR model by considering some important factors such as; the stress on the health care system, the financial loss, the activity level during lockdown, and individual behavior factor. Our contributions include the following epidemic management features:

- We investigate a state-space model using the real-time temporary sentiment to obtain the final sentiment information. We apply individuals’ posts on Twitter based on emotion and user interest that reflects public sentiment in real-time in regards to the current global outbreak.
- We consider an information system which is consisting of the processes of learning and forgetting of information. This model considers the impact of individuals’ propensities toward pandemic.
- In the proposed policy, population is divided into two groups based of individuals’ propensities. The percentage of individuals with high-risk behavior is *S*^*H*^, the percentage of individuals with low-risk behavior is *S*^*L*^.
- We suggest an online control strategy using data-driven method that provides a real-time health data such as the percentage of individuals who want to wear a mask *S*^*M*^.
- We propose an optimal lockdown policy that minimizes the financial loss which is associated with social performance and the stress on the health care system. The efficiency measures such as the speed of learning in the health care system during the pandemic are also considered in the proposed control policy.

The next section shows details of our proposed optimal control strategy that considers online disease information system to minimize the financial loss during the pandemic. In Section 3 we will show how are optimization model can be implied to real data. As an example we consider LA County. Section 4 summarizes our main findings and discusses possible directions for future study.

## 2 The proposed control strategy of pandemic

### 2.1 The disease information

Within a routine day each person makes both local and global communications. Each individual has a unique understanding from the epidemic and chooses different precautions to protect themselves from the virus. They receive information about disease from both their local and global networks. Local networks such as home or their neighborhoods, global network such as social media. Based on the amount of the disease information that individuals acquire they change their behavior during the epidemic.

Social media offer a platform for sharing disease information [9]. Currently people use the microblogging platform Twitter with over 300 million monthly users to share their ideas and feeling about a wide range of subjects. The governmental agencies such as the Centers for Disease Control and Prevention (CDC) and the World Health Organization (WHO) have used Twitter’s potential to update people about disease information. Using Twitter data to get an understanding of real-time individual’s behavioral response to an event is a well-accepted tool. For instance, during Ebola virus (EV) outbreak in 2014 [10], during the spread of influenza in 2009 [11], during Syndrome outbreak in 2015 [12] and the Zika virus epidemic [13] researchers used Twitter dataset. Tweets which demonstrate public sentiment in regarding with the outbreak are related to emotions of fear, anger and surprise. In this research, we extract samples from tweets which contain positive and negative attitudes about the current pandemic to determine the temporary public sentiment. We develop a statistical technique for predicting and tracking sentiment polarity and individuals’ behaviors from social media during health emergencies such as COVID-19. The disease information system is characterized through the definition of input or observed information, state variables, and output information [14]. The observed information are external entities that are added into the system and can serve as control noise or inputs. State variables are unobserved information that evolve through time following a given state equation and also depending on the values of the observed information. Lastly, output information results from the realization of the state plus noise factors and represent the observable outcome of the information system [15]. We run a state-space model which is a flexible framework with time-varying parameters to forecast the temporary sentiment polarity over time.

Suppose *y*_1_,…, *y*_*n*_ is a series of fear-sentiment observations at time *n* and *p*_1_,…, *p*_*n*_ refer to the unobserved states at time *n* that depend on the observation sets. The main idea behind state-space approach is to predict the state variables that depend on the observed sentiment information. The following equations show the observation equation and the state equation, respectively:

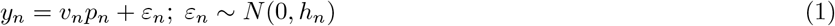

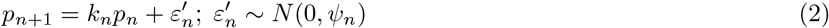

where the error terms *ε*_*n*_ and 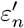 are assumed to be independent of each other. Let *ψ* _*n*_ and *h*_*n*_ denote the covariance matrices of the error terms, *v*_*n*_ and *k*_*n*_ are the ones that define how the observations relate to the state and how the state evolves over time.

Memory cells can store and retrieve the disease information. The disease information is being forgotten over time. Jaber and Bonney [16] studied the learning and forgetting information process. If *a* represents the degree of learning and *b* denotes the rate of forgetting of information, then after *n* time periods, memory performance, *z*, can be measured by the following formula.

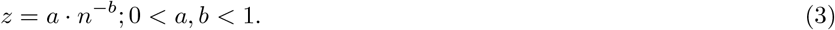

An individual is able to learn new information by *α*_0_ *p*_*n*+1_, while the past information is being forgotten by (1 − *α*_0_)*f*_*n*_. Finally, the final sentiment, *f*, is (Bi et al, [17]) calculated as follows,

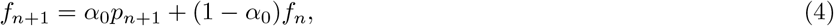

where 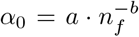 is the memory performance over the longest memory epoch *n*_*f*_.

### 2.2 The individual with low-risk behavior

When facing a pandemic different individuals have different responses. Some might self isolate most of the time, some might keep some of their social activities and some never follow any social distancing rules. In fact a wide range of rationale is involved. It could be because of the type of the job that a person has, like social workers, medical personnel and cab drivers, but here in this study we are considering the impact of fear sentiment. Behavior of individuals heavily depends on the amount of fear sentiment which they receive (Bi et al [17]).

As mentioned above, we denote the individuals with low-risk behavior by *S*^*L*^, where superscript *L* stands for low-risk. To calculate the fraction of population with low-risk behavior we use a “satellite” equation [18]. Jahedi and Yorke in [18] introduce the idea of using satellite equations. A satellite equation is an equation that is added to a model to calculate new quantities without changing the original model. We use the “Model E+” introduce by Jahedi and Yorke in [18] as our primary model to describe the pandemic. The Model *E*+ is as follows,

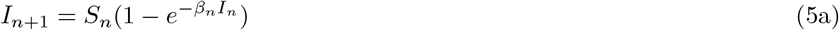

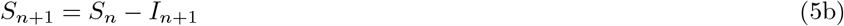

where *I*_*n*_ denotes the fraction of individuals who are infectious throughout the week *n* and *S*_*n*_ denotes the fraction of individuals who are susceptible at the beginning of week *n* respectively. The only parameter of the above model that needs to be specified to produce the pandemic is the contact rate in week n which is denoted by *β*_*n*_. According to Poisson distribution when the expected number of events in a unit time is *λ* and events occur independently, then the probability that an event does not occur is *exp*(−*λ*). Susceptible individuals on average make *β*_*n*_*I*_*n*_ infectious contact, therefore the probability that no new infectious case happens in week *n* is *exp*(−*β*_*n*_*I*_*n*_), therefore the number of new infectious cases in week *n* + 1 will be calculated by Eq. 5a above.

Let us assume that we want to produce the pandemic using the real data. By assuming that *S*_0_ = 1, i.e. initially everyone is susceptible, then Model E+ (5) suggests that we can calculate the contact rate in week *n* by the following equation.

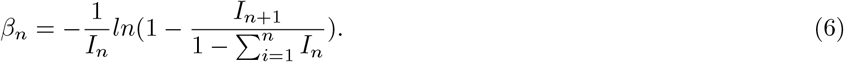

As mentioned above *I*_*n*_ is the fraction infectious in week *n* and can be calculated using real data.

Once we calculate the contact rates using Eq. (6), we can plug it in Model (5) and produce the outbreak. Now we have the fraction susceptible in each week, we finally calculate the probability that an individual wants to have low-risk behavior by the next satellite equation Chen et al [19],

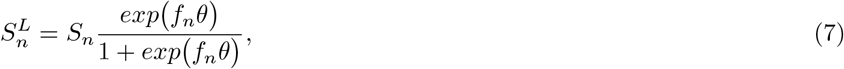

where *θ* is a coefficient to amplify the final sentiment and 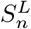 is the percentage of susceptible individuals with low-risk behavior. The ratio *exp*(*f*_*n*_*θ*)/(1 + *exp*(*f*_*n*_*θ*)) is the rate at time period *n* at which susceptible individuals choose to have low-risk behaviors.

#### 2.2.1 Why we should all be wearing face masks

COVID-19 is a global pandemic that mostly spreads by asymptomatic or presymptomatic patients because individuals often do not know if they are infected with the COVID-19. Therefore, it is difficult to determine the percentage of infected individuals by the classical SIR model when almost everyone is susceptible. To address the issues, we calculate the susceptible population based on individuals’ propensities that considers the susceptible individuals with high-risk and low-risk behaviors.

Face masks combined with other preventive measures, such as self-quarantine and frequent hand-washing, help slow the spread of the virus [20]. The Disease Control and Prevention (CDC) recommends community face masks for the public, while the N95 and surgical masks are needed by health care providers. Modeling and tracking public sentiment can provide key information for decision making to control the pandemic. Using the temporary public sentiment related to masks, we calculate the probability that an individual wants to wear mask by the next satellite equation,

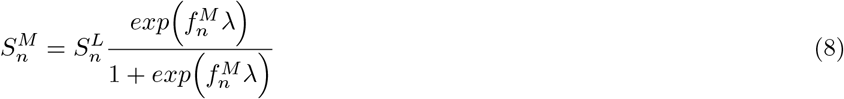

where 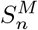 is the ratio of individuals with low risk behaviors who wear mask and 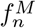 denotes the final sentiment related to masks and *λ* is a coefficient that amplifies the public sentiment polarity about masks.

### 2.3 Optimal threshold between health care system and economic activity

Another major open question, affecting the decisions of policy makers, is how to calculate the relation between health care system and economic activity using the actual number of COVID-19 infections. To address this issue, we introduce an optimization formulation to obtain the optimal activity level during lockdown that minimizes the financial loss and the stress of health care system. The first term in the right hand side of Eq. (9a) is the stress in the health care system that determines by proportional to 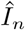 a factor *c*_2_ that is the efficiency level or the speed of learning related to the health care sector, and a coefficient *c*_1_ that shows the burden of new infections on the health care system which is calibrated by the damage of the pandemic on economic about the U.S. of 13 trillion dollars corresponding to 61% of the annual U.S. GDP (Gonzalez [21]). The relation between the activity level (social performance) and financial loss during the pandemic is considered in the second term in the right hand side of Eq. (9a) where *ω*_1_ and *ω*_2_ are the scaling and shift parameter (Chen et al. [19]).

The constraint Eq. (9b) is a possibly time-varying infection rate that reflects government policy such as the activity level. By increasing activity level 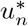 the percentage of infection goes up that causes a burden on the health care sector. Let *γ* denote the growth rate of the percentage of infected individuals [10, 22]. The last constraint is the optimal activity level 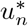 which is limited between 0 and 1.

**Problem 1. The proposed optimal strategy for the COVID-19 pandemic**

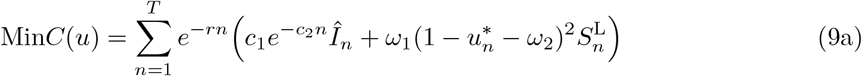

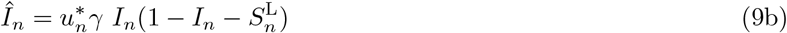

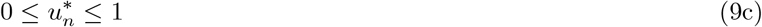

Using the proposed algorithm (which is called Algorithm 1), by forecasting the temporary public sentiment, first we calculate the final sentiment (FS) using the rate of learning and forgetting information. We determine the percentage of infected persons and the percentage of susceptible persons using the SIR model. Then, we determine the percentage of individuals who have high fear and follow low-risk behavior. The percentage of people who wear a mask is also determined by a satellite equation. Finally, we solve an optimization model to find optimal threshold of activity level and the financial loss during the epidemic.

#### Algorithm 1. Proposed optimal control algorithm for COVID-19pandemic

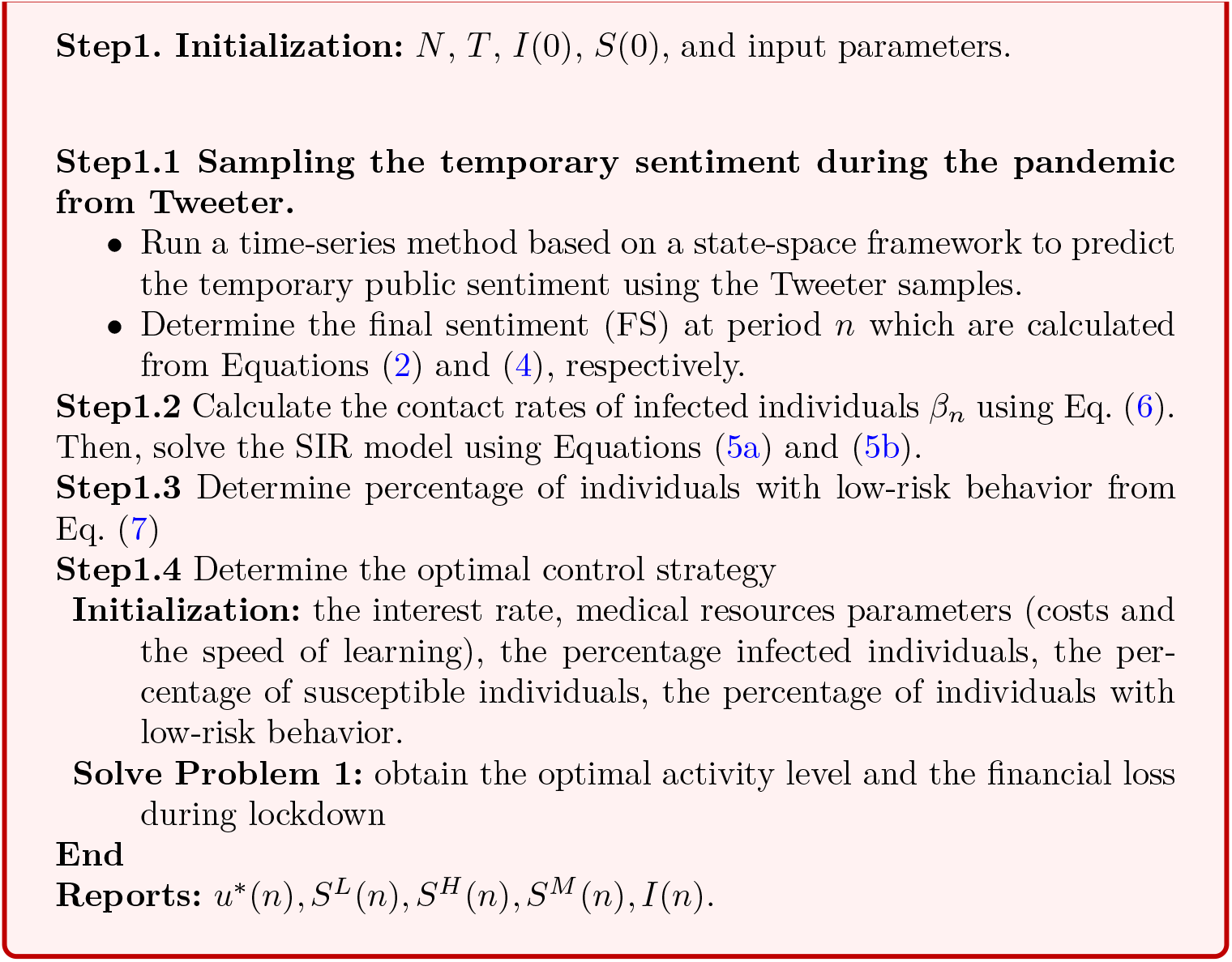

## 3 Experimental results

### 3.1 COVID-19 dataset

Twitter is a valuable platform for analyzing and tracking public sentiment where millions of users share their feelings. We carried out an experimental study of the proposed model by Tweet dataset in California. We are continuously gathering the dataset since March 5, 2020 until July 6, 2020. The data that were obtained from [23] were used to evaluate the framework. Each file has about 200 thousand rows, and each row contains tweet id, date/time, location, text, user id, and user verified. The polarity of the sentiments is distributed across the scale between [-1,0), 0, and (0,+1] that shows negative, natural, and positive polarity, respectively. We extract samples from tweets which contain positive and negative attitudes about the current pandemic to determine the temporary sentiment information. Figure 1 represents the number of neutral, negative, and positive tweets. Other datasets related to the COVID-19 pandemic such as the number of confirmed cases in LA County can be also found in the USAFacts dataset [24].

**Figure 1:**
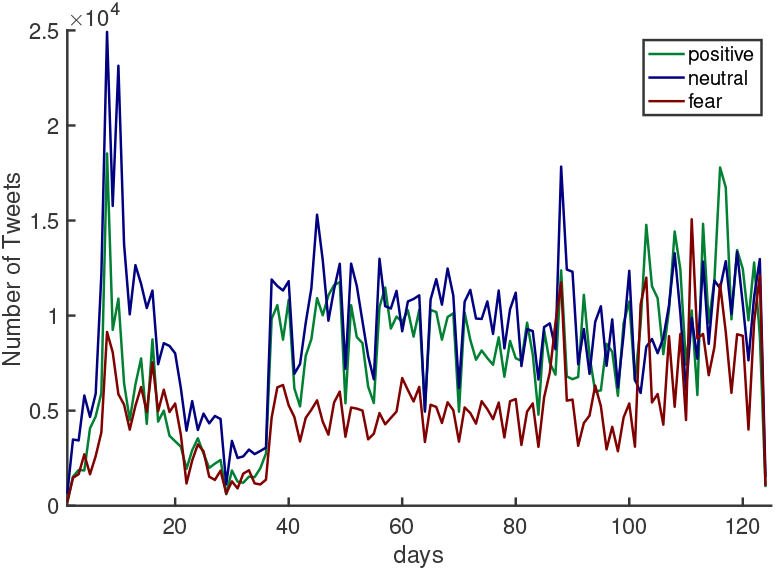
Temporary sentiment from March 5th to July 6st, 2020 (Source: Twitter)

We run a time-series analysis to predict the range of temporary sentiment over time. Figure 2 predicts the temporary sentiment information using time-series method in a state-space framework.

**Figure 2:**
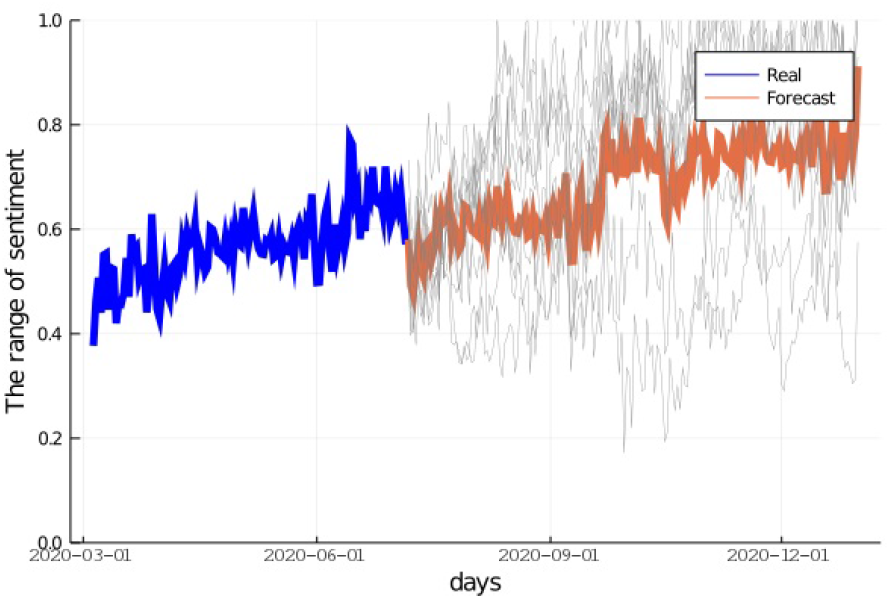
Historical temporary sentiment and forecasting over 100 simulated runs

### 3.2 Results

#### 3.2.1 Optimal control evaluation of disease

The list of input parameters used for the SIR model is presented in Table 1. Using the range of temporary sentiment, we solve a SIR model to determine the percentage of infected individuals *I*(*n*), the percentage of susceptible individuals with high risk behaviors *S*^*H*^ (*n*), the percentage of susceptible individuals with low-risk behavior *S*^*L*^(*n*), and the percentage of susceptible individuals with low-risk behavior who wear mask *S*^*M*^ (*n*). Lastly, we obtain the optimal activity level that minimizes the financial loss during the pandemic. Figure 3 presents the contact rate of detectable infected individuals over time in LA County which is calculated using Eq. (6).

**Table 1:** The model parameters

| Parameter | Description | Value |
| --- | --- | --- |
| $\theta$ | coefficient to amplify the FS | 1 |
| $a$ | the degree of learning | 0.5 |
| $b$ | the rate of forgetting information | 0.5 |
| $n_f$ | the sum of memory portions | 7 days |
| $c_1$ | a coefficient (the burden of new infections on the health care sector) | 265 ([21]) |
| $c_2$ | the speed of learning for the health care service | $-\ln(0.5)/182$ ([21]) |
| $r$ | an interest rate | 0.0001 |
| $\omega_1, \omega_2$ | scaling and shift parameter for financial loss performance | 0.2, 0.3 |
| $\lambda$ | coefficient to amplify the FS related to masks | 10 |
| $\gamma$ | the growth rate of infection | 0.5 |
| $I(0)$ | initial infection rate | $1/N$ |
| $S(0)$ | initial susceptible rate | $(N - 1)/N$ |
| $N$ | population(LA County) | 10,000,000 |

**Figure 3:**
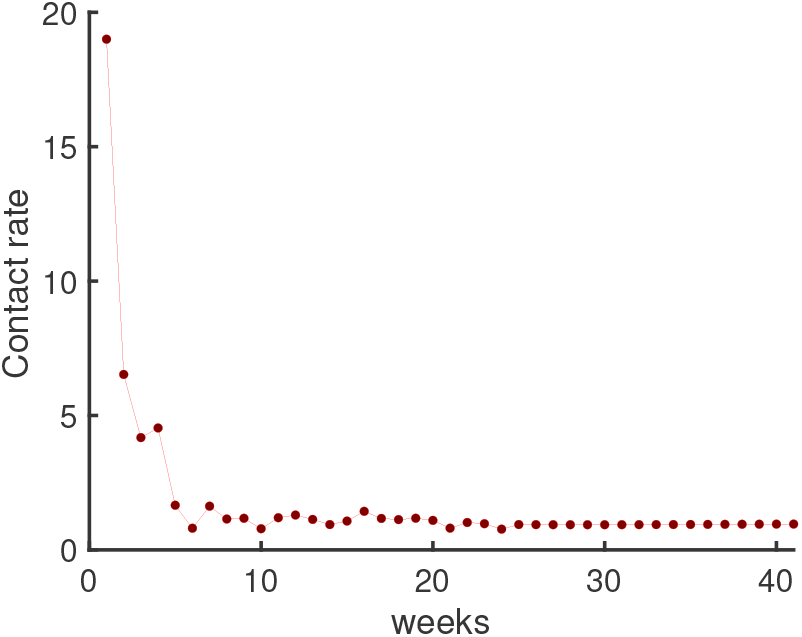
The contact rates of detectable infected individuals over time in LA County

We now can determine the number of susceptible and infected individuals using Model E+ 5. The number of infected individuals in LA County over 40 weeks is shown in Figure 4.

**Figure 4:**
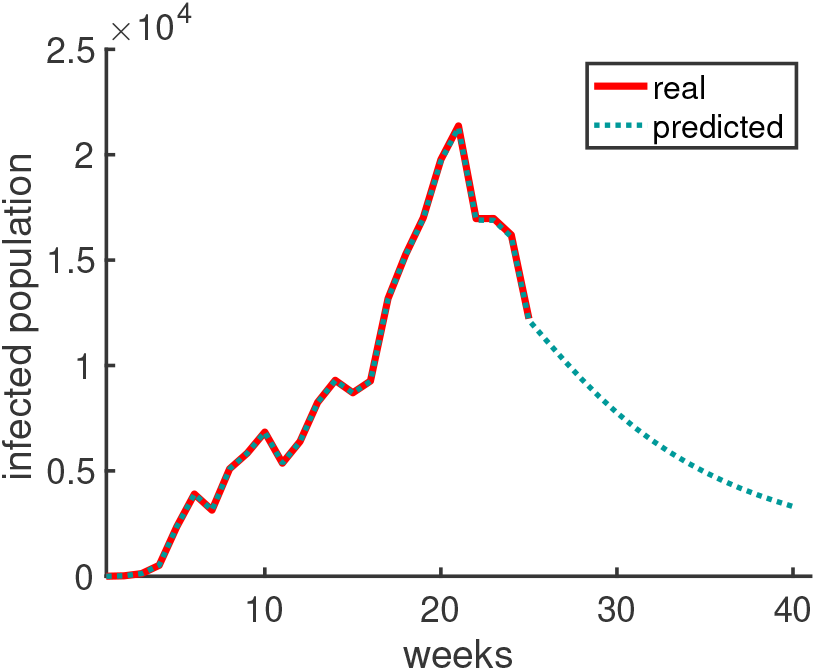
The number of infected individuals over time in LA County

In this study, we consider the susceptible individuals under two types of behaviors. One group is the percentage of susceptible individuals with risky behaviors, while other group is the percentage of susceptible individuals with low-risk behaviors. Individuals will change their behaviors during the pandemic based on the final sentiment information which they receive. The total number of susceptible individuals is shown in Figure 5a and the number of susceptibles with low-risk behavior is shown in Figure 5b.

**Figure 5:**
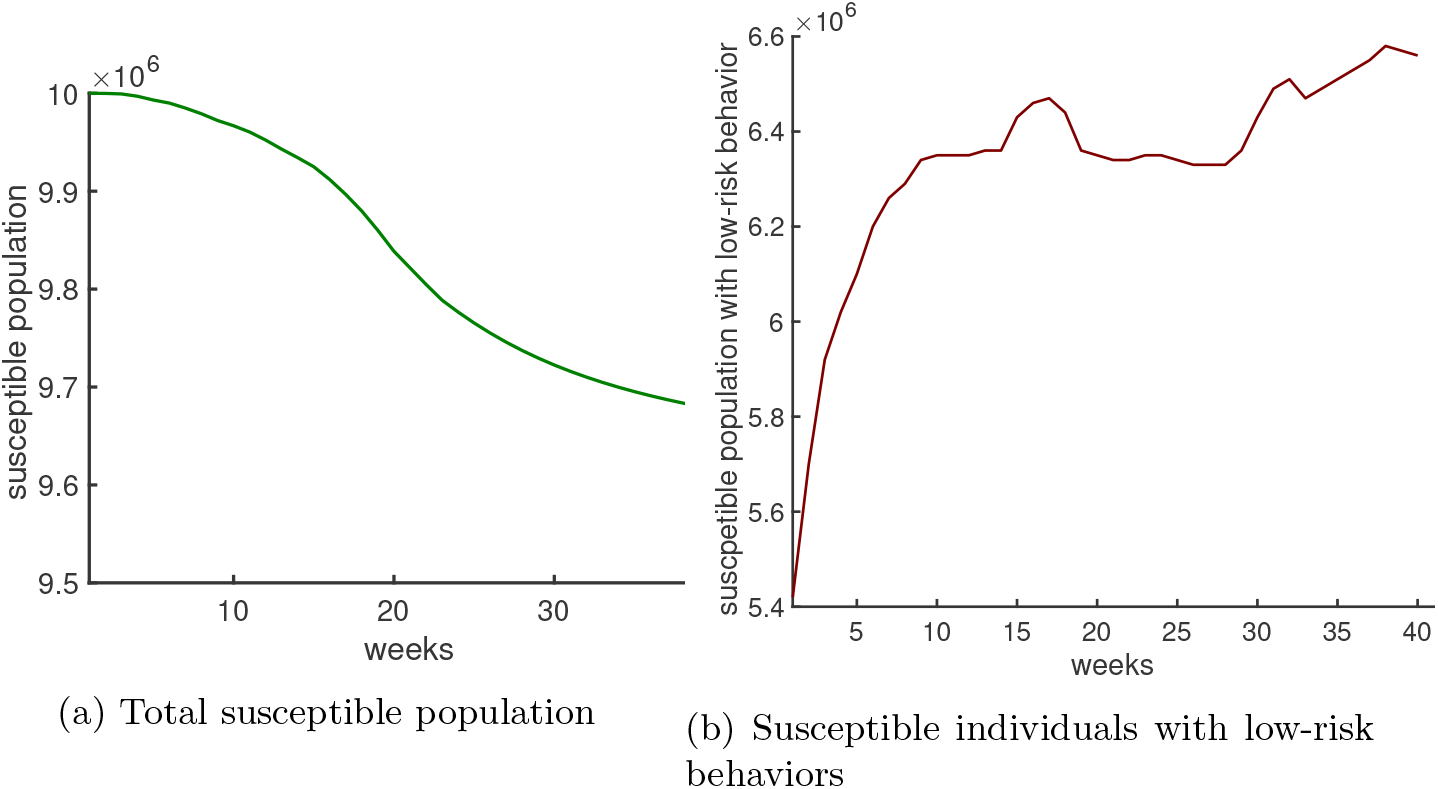
Susceptible individuals

The percentage of individuals with low-risk behaviors can help the service providers to find the optimal response and recovery strategy during the epidemic. For example, the transit system needs to estimate the actual demands and design the fleet during the epidemic. Additionally, suppliers can take advantage of the activity level information to manage supply chain risk and disruption in directing the flow of goods to demand nodes during the pandemic. As Figure 5b represents the percentage of susceptible with low-risk behaviors increases at the first of the epidemic, because the individuals have high fear. However, it goes down at the peak of the COVID-19 pandemic and finally it increases during the remaining time periods of the pandemic.

The health care sector and medical professionals are facing challenges like never before due to the COVID-19 pandemic. We solve an optimization model based on Eq (9) that minimizes the total cost in terms of the stress of health care system and the financial loss associated with social performance. Prediction of the pandemic duration can help the policy makers to forecast the end time of lockdown to avoid consequent social-economic damages as well. The optimal activity level during lockdown is determined using the provided information by solving the SIR model. A 3.2 trillion dollars gross state product as of 2019 is showed in the California state that uses to estimate the percentage damage during the pandemic [25].

This study explores the determinants of switching behaviors in the social activity level. The impact of heterogeneous susceptible (who have low-risk behavior *S*^*L*^ and wear a mask *S*^*M*^) on 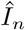 is also considered in Eq. (9b). Figure 6a shows the impact of individual behaviors on the optimal activity level during the epidemic in LA County. The results show that the optimal activity level is decreased until 0 % during weeks 18 through 24 where a homogeneous population of susceptible is considered. The social performance level is dropped until 40 % when considering a heterogeneous population of susceptible that includes individuals with high fear. The impact of susceptible individuals who wear a mask is also shown in Figure 6a, while the optimal activity level is reduced until until 10 %. Likewise, a sensitivity analysis on *γ* (between 0.1 and 0.7) and the optimal activity level that reflects the stress of health care system during the pandemic is shown in Figure 6b.

**Figure 6:**
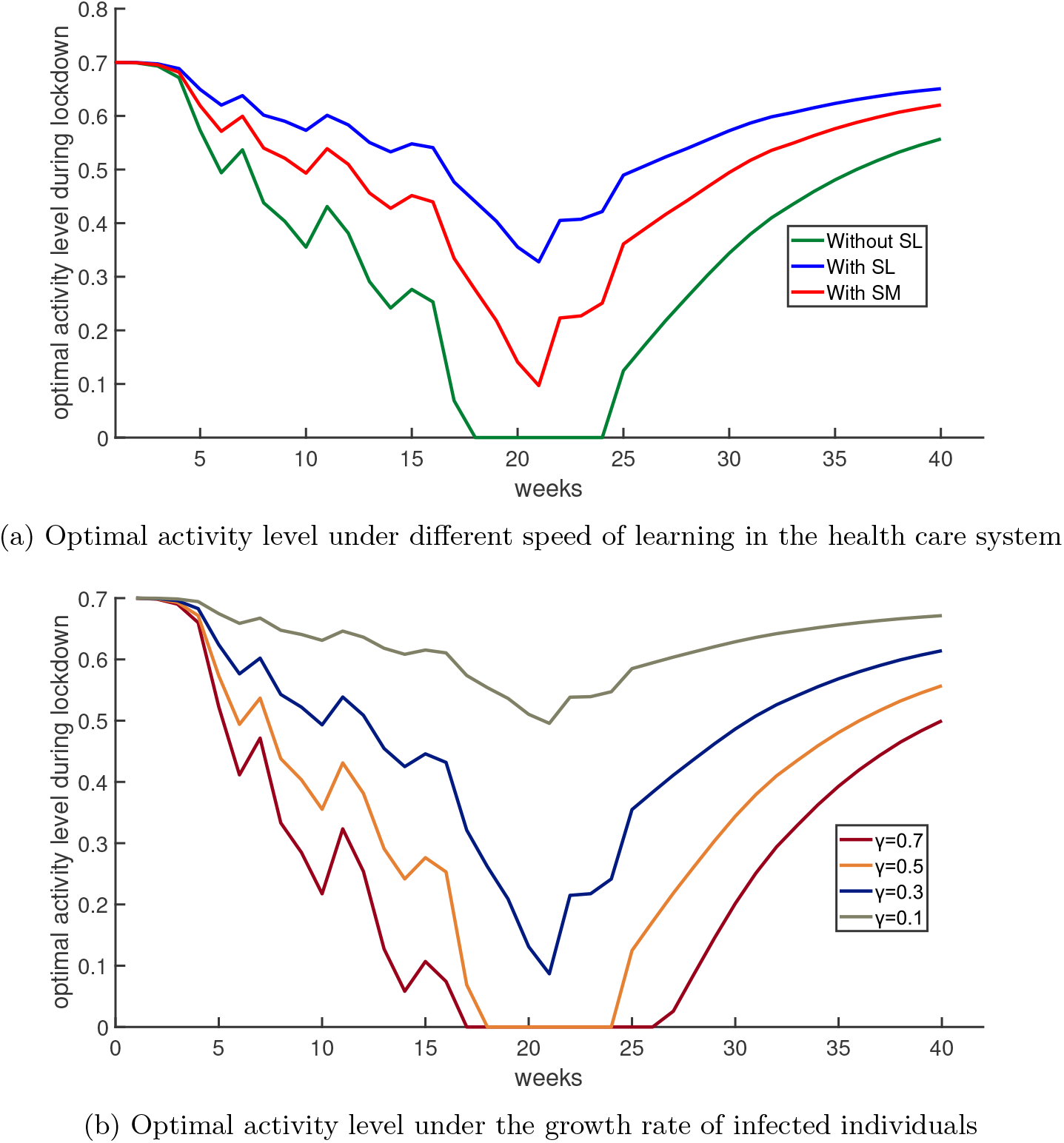
Sensitivity analysis

#### 3.2.2 Online social distancing monitoring

Wearing a mask helps prevent the spread of COVID-19, especially those at highrisk. In this study, we use the tweets about mask to analyze the effect of face masks on the percentage of susceptible people who follow social distancing. The number of tweets related to masks in California from March 5*th* to July 6*th*, 2020 is 86365. Daily number of tweets related to masks is shown in Figure 7a. Before May 30, 2020, our analysis showed a very small number of tweets related to mask followed by a steady increase starting June 1, 2020. We analyzed tweets about masks for negative, neutral, and positive polarity. Figure 7b shows an analysis of tweets sentiment polarity about masks. Using the temporary sentiment related to masks, we are able to calculate the percentage of individuals who want to wear mask. Figure 8 shows susceptible individuals with low-risk behavior and susceptible persons who wear a mask. Analysis of such data showed that 77% of people in LA County wear mask; this agrees with the results of [26, 27]. CDC investigated dataset from an internet survey of a national sample of 503 adults and the result indicates that the percentage of adults endorsing face mask wearing is about 76% [26]. Therefore, not only the proposed online strategy mitigates the pandemic, but also provides real-time health data based on individuals’ propensities.

**Figure 7:**
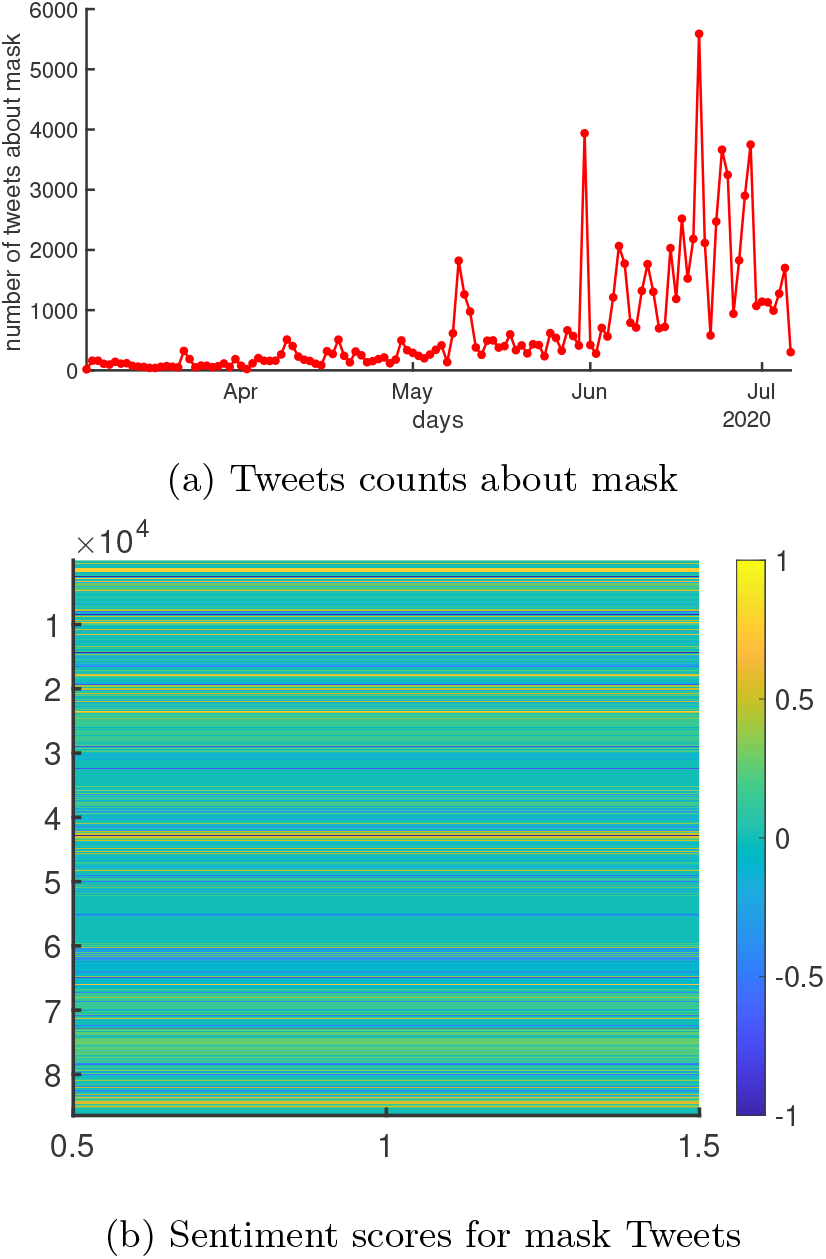
Tweets related to mask from March 5th to July 6st, 2020

**Figure 8:**
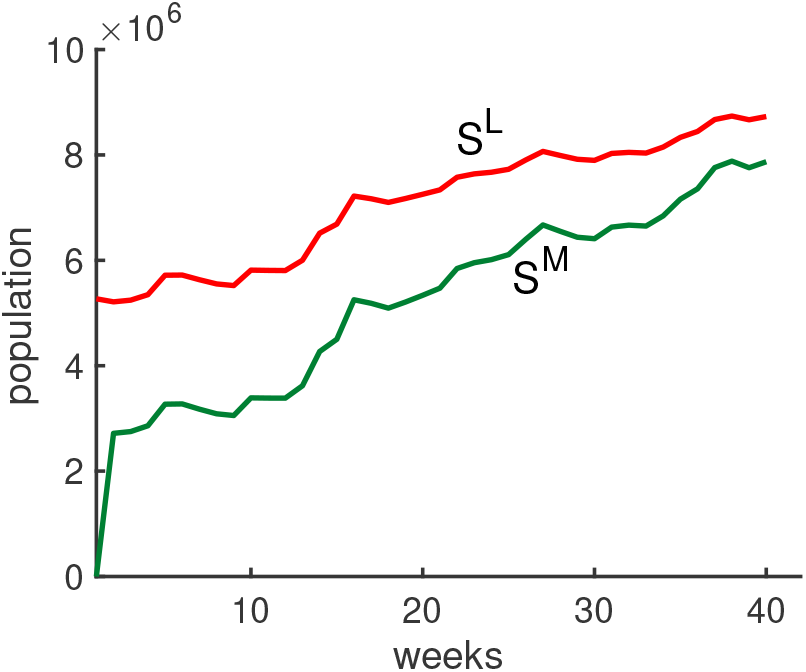
Susceptible individuals with low-risk behavior and susceptible individuals who wear a mask

#### 3.2.3 The sensitivity analysis of learning and forgetting disease information

The individuals communicate by sharing their sentiment to each other where their individual’s brains store and retrieve the disease information during the pandemic. The individual’s brain can make lots of daily decisions about whether to store facts and events. Some of experiences will be stored in the brain for a few seconds or minutes and finally forgotten. However, some of experiences and facts will be remained for a few days, while others will be ingrained for many years or even a lifetime. If individuals decide to remember the disease information, the brain makes connections between the cells, which alters their structure, and is what allows individuals to retain memories. We determine the final sentiment (FS) with regards to the learning and forgetting factors for individuals.

We calculate the individual’s memory performance measure based on Eq. (3). A sensitivity analysis of the FS by changing the forgetting factor of individual’s memory between 0.1 and 0.9 is shown in Figure 9a. As a result, the rate of the FS goes up by decreasing the rate of forgetting information from 0.1 to 0.9. Similarly, we analyze the relation between the longest memory epoch *n*_*f*_ and the final sentiment (FS) as shown in Figure 9b. We compare the number of susceptible individuals with high fear by changing the rate of forgetting information *b* between 0.1 and 0.9.

**Figure 9:**
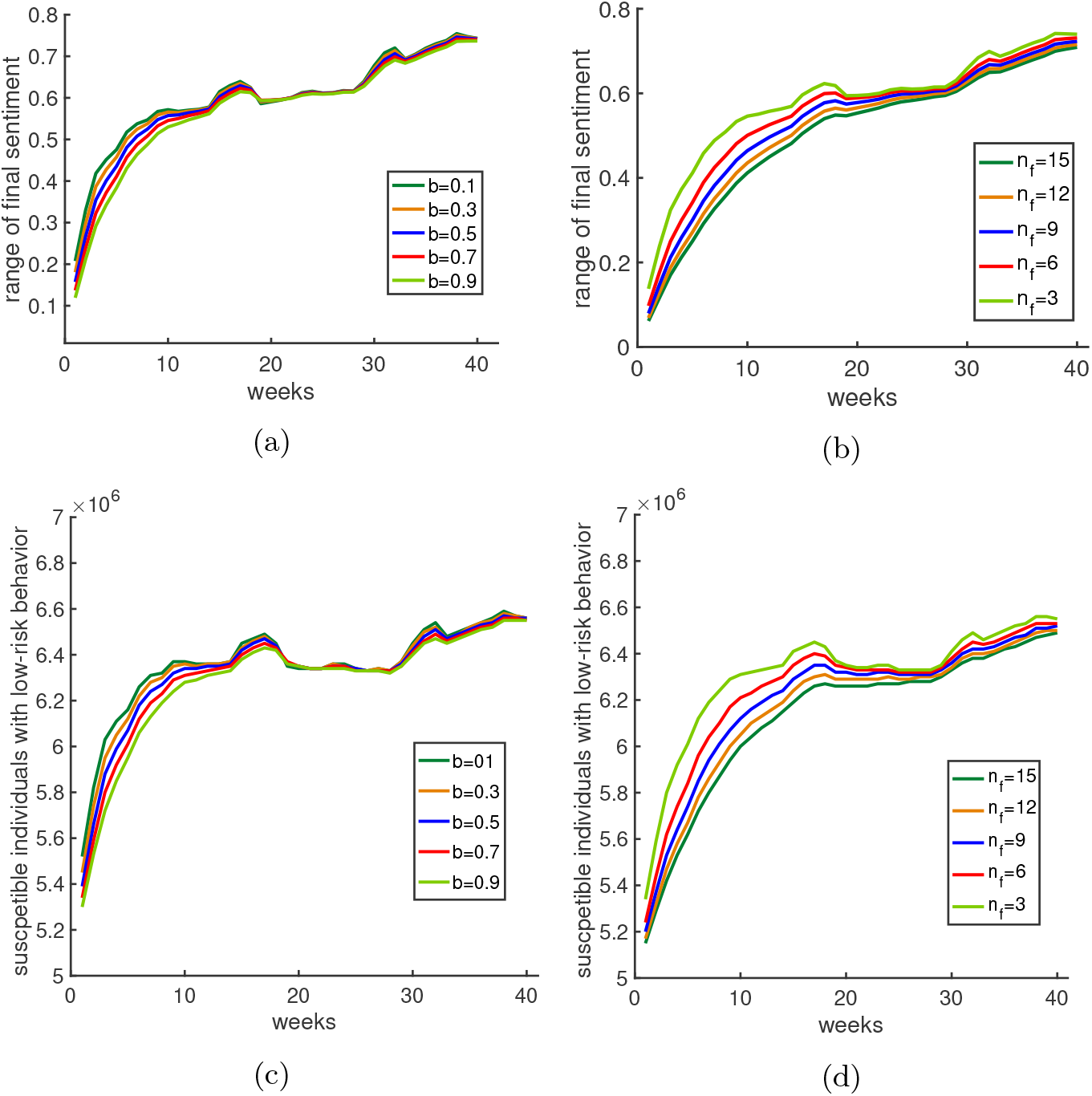
(a) Final sentiment (FS) under different rate of forgetting information(LA County). (b) A sensitivity analysis on the longest memory epoch *n*_*f*_ and the final sentiment (FS). (c) A sensitivity analysis on the forgetting sentiment information and the susceptible individuals with high fear. (d) A sensitivity analysis on the longest memory epoch *n*_*f*_ and the susceptible individuals with high fear.

Figure 9c shows a sensitivity analysis on the forgetting sentiment information and the number of individuals who have low-risk behaviors. The low-risk behaviors during the pandemic has also affected by the longest memory epoch *n*_*f*_. Thus, we analysis the relation between the longest memory epoch *n*_*f*_ and the number of susceptible individuals who have high fear as shown in Figure 9d. Individuals will have the lower risk behavior and higher infections when the longest memory epoch *n*_*f*_ goes up.

## 4 Conclusions

This paper presents the impacts of the COVID-19 pandemic using the complex interactions among disease, social and economic systems and infrastructures. We proposed an optimal strategy model for pandemics that considers the interaction of individuals to obtain the temporary sentiment information and the final sentiment information. A fear information function based on the final sentiment information is calculated to find the percentage susceptible individuals with high fear and low fear. Then, we proposed the SIR model for COVID-19 to determine the percentage of infected persons and the percentage of susceptible persons over time. Lastly, we solved an optimization formulation to find trade-off between social performance and limited medical resources. We proposed optimal lockdown and exit strategy during an epidemic for each state or territory that helps the policy makers during the pandemic.

The proposed model helps us to understand the impact of epidemic on transport modes and travel patterns of individuals. Such this information helps us to solve the challenges of the last mile delivery, multi-tiered delivery, and delivery on demand during the pandemic. For example, the transit service providers can control customer demands during the pandemic by predicting the percentage of individuals who have participated social distancing. Additionally, the proposed policy can monitor the human behaviors during epidemic in order to control the stress of health care sector. The proposed optimal strategy also provides a systematic recovery strategy after the pandemic. Other human trajectory datasets collected from the movement of individuals during the pandemic can be used to update the proposed optimal control strategy model [28]. Finally, an agent-based simulation based on the human trajectory for estimating the transmission of disease information between agents during the pandemic would be highly valuable.

## Data Availability

Upon request all data will be provided.

## Acknowledgments

The author appreciates helpful comments from Sana Jahedi at University of New Brunswick. This research did not receive any grant from funding agencies in the public, commercial, or not-for-profit sectors.

## Authors’ contributions

Our contributions include the following epidemic management features: When an outbreak starts spreading, policy makers have to make decisions that affects health of their citizens and the economic. Some might induce harsh measures, such as lockdown. Following a long harsh lockdown, economical declines force policy makers to rethink reopening. But what is the most effective reopening strategy? In order to provide an effective strategy, here we propose a control strategy model. Our model assesses the trade-off between social performance and limited medical resources by determining individuals’ propensities. The proposed strategy also helps decision makers to find optimal lockdown and exit strategy for each region. Moreover, the financial loss is minimized. In addition, a study of a COVID-19 dataset for Los Angeles County is performed to validate our model and its results. Our contributions include the following epidemic management features:

Department of Systems Engineering, Cornell University, Ithaca, USA.

